# Research Priorities of Individuals and Families with Sex Chromosome Aneuploidies

**DOI:** 10.1101/2024.08.15.24312069

**Authors:** Alexandra Carl, Samantha Bothwell, Fathia Farah, Karli Swenson, David Hong, Siddharth Prakash, John Strang, Nicole Tartaglia, Armin Raznahan, Judith Ross, INSIGHTS Consortium, GALAXY Consortium, Shanlee Davis

## Abstract

Sex chromosome aneuploidies (SCAs) are chromosomal variations that result from an atypical number of X and/or Y chromosomes. Combined, SCAs affect ~1/400 live births, including individuals with Klinefelter syndrome (47,XXY), Turner syndrome (45,X and variants), Double Y syndrome (47,XYY), Trisomy X (47,XXX), and rarer tetrasomies and pentasomies. Individuals with SCAs experience a wide variety of physical health, mental health, and healthcare experiences that differ from the standard population. To understand the priorities of the SCA community we surveyed participants in two large SCA registries, the Inspiring New Science in Guiding Healthcare in Turner Syndrome (INSIGHTS) Registry and the Generating Advancements in Longitudinal Analysis in X and Y Variations (GALAXY) Registry. 303/629 (48.1% response rate) individuals from 13 sites across the United States responded to the survey, including 251 caregivers and 52 self-advocates, with a range of ages from 3 weeks to 73 years old and represented SCAs including Turner syndrome, XXX, XXY, XYY, XXYY, and combined rare tetrasomies and pentasomies. Results demonstrate the priorities for physical health and emotional/behavioral health identified by the SCA community, as well as preferred types of research. All SCA subtypes indicated intervention studies as the top priority, emphasizing the need for researchers to focus on clinical treatments in response to priorities of the SCA community.

**STATEMENTS RELATING TO ETHICS AND INTEGRITY POLICIES:** The data that support the findings of this study are available on request from the corresponding author. This study was funded by the Turner Syndrome Global Alliance, Association for X and Y Chromosome Variations, Living with XXY, the XXYY Project, and the University of Colorado School of Medicine Department of Pediatrics. Data collection and storage was supported by NIH/NCATS Colorado CTSA Grant Number UM1 TR004399. The authors do not have any conflicts of interest to disclose. This study was reviewed and approved by the Colorado Multiple Institutional Review Board (COMIRB # 20-0482 and # 19-3027). All participants provided informed consent for participation - participants under 18 years old provided assent along with parental informed consent prior to any study procedures. Contents are the authors’ sole responsibility and do not necessarily represent official NIH views.

## 1. INTRODUCTION

Sex chromosome aneuploidies (SCAs) are a set of rare genetic conditions in which there is either a missing or additional sex chromosome(s). These include Klinefelter syndrome (KS, 47,XXY; 1 in 600 male births^1^; Double Y syndrome (47,XYY; 1 in 1,000 male births^2^; Turner syndrome (TS, 45,X and variants; 1 in 2000 live female births^3^); Trisomy X (47,XXX; 1 in 1,000 live female births); and other much less common sex chromosome tetrasomy and pentasomy conditions. The phenotypic characteristics of SCAs vary widely across different variations and among individuals with the same diagnosis. Individuals with SCAs may present with various developmental, neurocognitive, or psychological challenges, as well as myriad physical and reproductive health challenges^4–7^.

Historically, in clinical research, physicians, scientists, advocacy groups, and funding agencies have influenced research priorities to advance their respective specific objectives^8^. Since deficits in knowledge continue to exist in SCA care, it is crucial to consider community priorities when determining future directions for research. As a result, past research has often been confined to single sites with limited participant diversity or focused on specific features (e.g., growth in TS), with most prior research focused on physical conditions. Research resulting from this prioritization method has not fully addressed all clinical needs, and deficits of knowledge continue to exist in SCA care.

One prior study by Sandberg et al in 2019 surveyed the TS community and found that patients and families place importance on both physical and emotional health research; however, most research has focused on physical conditions^9^. While these findings were important in highlighting the need for research on psychological features of TS, it did not probe on specific topics, but rather asked participants about the topic areas of “medical/physical problems,” “emotional/behavioral problems,” and “participating in Turner syndrome research.” Within the latter category, participants were specifically asked about five areas: “new medicines,” “medical devices,” “eating or nutrition,” “quality of life,” and “genetics.” This emphasizes the need to understand more granular research priorities of this population. Studies exploring the research priorities of patients and families in other SCA conditions are lacking. Thus, there remains an unmet need to understand granular research priorities of patients and families in all SCA conditions.

This study utilized community-engagement methods with support from the stakeholder steering committees for the Inspiring New Science in Guiding Health Care in Turner Syndrome (INSIGHTS) and Generating Advancements with Longitudinal Analysis in X and Y Variations (GALAXY) Registries, to create surveys tailored to the SCA population^10,11^. The goal of this study was to determine the health areas of highest research priority to the SCA community, further exploring differences in priorities when stratified by SCA group and when compared by respondent (self-advocate versus caregiver).

## 2. MATERIALS AND METHODS

### 2a. Participant recruitment

Data analyzed for this report were derived from the GALAXY and INSIGHTS projects^12^, which collect clinically verified and self-report data from individuals with SCAs evaluated at 13 TS or SCA multidisciplinary clinics across the United States. Enrollment criteria included a genetically confirmed SCA diagnosis, which is validated for each participant by study staff. These analyses were conducted on the cross-sectional baseline dataset from both databases. Data were analyzed on all individuals with 1.) values for basic demographics and 2.) complete responses to the Research Priorities survey.

Individuals with an SCA (referred to henceforth as self-advocates) or caregivers of individuals with SCA (referred to henceforth as caregivers) could respond to the survey. The survey was completed at home using the online REDCap platform^12^ or in clinic on an iPad after enrollment into the study, and respondents included here completed the survey between February 23, 2023 and July 2, 2024.

#### Editorial policies and ethical considerations

The Colorado Multiple Institutional Review Board (IRB) reviewed and approved both studies, acting as the single IRB for all sites (COMIRB #19-3027 for INSIGHTS and COMIRB #20-0482 for GALAXY). The study team obtained informed consent from participants over 18-years of age, and assent from participants younger than 18 years along with parental consent.

### 2b. Survey design

Both surveys were designed and modified by the Steering Committee (SC) for each registry to be inclusive of the range of health topics that are relevant to individuals with SCAs and caregivers. The SCs are comprised of clinical experts, researchers, individuals with SCAs, and caregivers of individuals with SCAs, and is designed to bring together a variety of expertise, experience, and opinions. SC members contributed additional relevant survey topics and edited the surveys for clarity and acceptability.

Priority for several health topics and research study types were measured with a slider scale ranging from 0 to 100, with 0 indicating the lowest priority and 100 the highest priority. Participants were instructed to, “indicate how [they] would like to see research on this area prioritized for the X&Y community. Think about what is important to [them] and what [they] want resources to go toward.” Participants were also instructed that, although they may feel everything is important, to use both sides of the scale so we can tell which topics were of higher priority than others (full instructions in Appendix 1). Topic areas were broken into four categories: medical/physical health (28 topics), emotional/behavioral health (11 topics), other (15 topics), and research study types (6 topics), with minor variations between the INSIGHTS (TS only) and GALAXY (all other SCA) surveys (Appendix 1).

### 2c. Statistical Analysis

Descriptive statistics are stratified by SCA condition and presented for demographic variables as N (%) for categorical variables and median [IQR] for continuous variables. Age reflects the age of the individual with an SCA at the date of survey response. Research priorities for each SCA condition are visualized as boxplots, depicting median [IQR]. Within each research domain (physical health, types of studies, emotional/behavioral health, and other health), research priorities are shown for the top 10 topics, given the median is greater than 50. Within domains that contained fewer than 10 research topics, all topics with a median greater than 50 are plotted. Overall self-advocate versus caregiver, as well as INSIGHTS versus GALAXY, reported scores are summarized as median [IQR] and differences were tested using Kruskal-Wallis tests and are interpreted under the assumption of a type 1 error rate of 0.05. Additional False Discovery Rate (FDR) adjusted p-values are presented to rule out truly null results. Comparison of research priorities are visualized using heatmaps, with the heatmap.2 R function within the gplots package, with columns and rows sorted by hierarchical clustering using Euclidean distance. All analyses were performed in R, version 4.4.0.

## 3. RESULTS

### 3a. Participant Demographics

A total of 303 of 629 (48.1%) participants (251 caregivers and 52 self-advocates) completed the survey (Table 1). Self-advocate participants were a median of 35.1 years old with a range from 13.4 to 73.8 years. The children of caregiver respondents were a median of 7.7 years old with a range from 3 weeks to 35.7 years. More than three-fourths (79.9%) of respondents identified as White Non-Hispanic and most participants (59.5%) held private insurance. Respondents represented ten multidisciplinary clinics across the United States, with the greatest (40.8%) from Children’s Hospital Colorado (CHCO). The sample was stratified by SCA diagnosis, with the largest cohort diagnosed with TS (n=144), followed by KS (n=86), XXYY syndrome (n=35), and Trisomy X (n=17).

### 3b. Research Prioritization by SCA

The top priorities overall in the TS participant group (Figure 1, Supplemental Table 1) were anxiety disorders (82 [67, 97.5]), fertility concerns (82 [68.5, 99]), and social deficits (80 [65, 96]). The top priorities in the Trisomy X group (Figure 2) were learning disabilities (96 [71, 100]), medical and healthcare access (89 [66.5, 100]), and, similar to the TS group, anxiety disorders (88 [75, 100]). The top priorities overall in the KS group (Figure 3) were similar to the TS group as well: hormone replacement therapy (94.5 [75, 100]), fertility concerns (88.5 [69.25, 100]), and learning disabilities (82 [67, 98]) tied with strengths-based research (82 [69.75, 98]). The top priorities overall in the XYY group, all caregiver respondents (Figure 4), were fertility concerns (100 [59.5, 100]), learning disabilities (98.5 [72.5, 100]), and self-harm/suicidality (93.5 [71.5, 100]). The top priorities in the XXYY group, all caregiver respondents again (Figure 5), were social deficits (91 [73, 98.5]), quality of life (87 [77.5, 98]), and hormone replacement therapy (85 [75, 97.5]) tied with self-sufficiency (85 [76, 98). The *all other SCA group* included those with 48,XXXY and pentasomies, and all but one were caregiver respondents. The top priorities overall in this group (Figure 6) were social deficits (85 [65, 92]), understanding X and Y variations (80 [70, 85]), and learning disabilities (78 [72, 95]) tied with self-sufficiency (78 [67, 97]).

**Figure 1.**
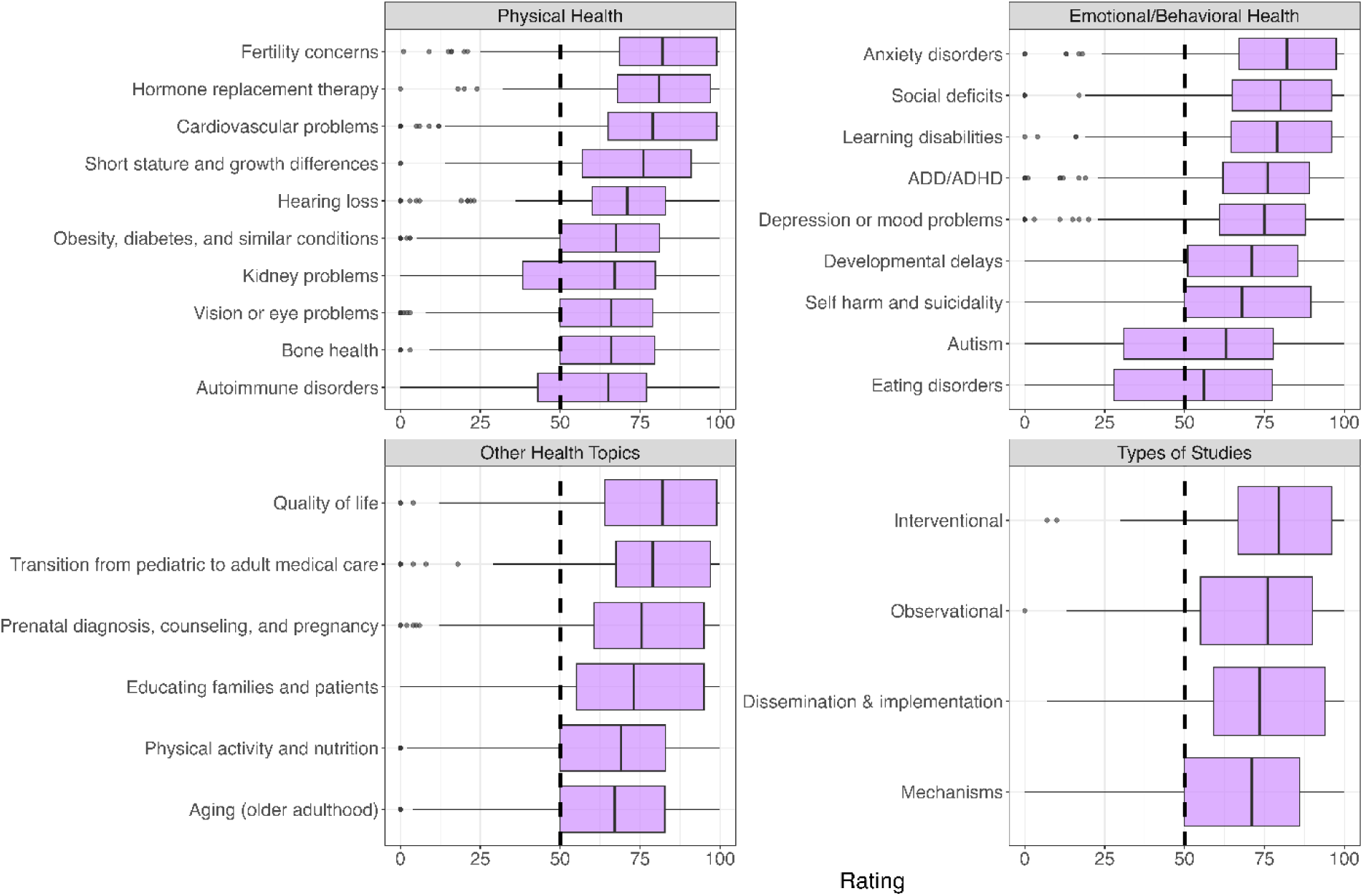
Turner syndrome research priorities. Research priorities among the Turner syndrome sample (n=144), including physical health, emotional/behavioral health, and other health domains, and types of research studies. Topics with a median greater than 50 are presented, capped at the top 10 topics per domain. The top physical health priorities reported by this group were fertility concerns (median [IQR] : 82 [68.5, 99]), hormone replacement therapy (81, [68, 97]), and cardiovascular problems 79 [65, 99]. The top emotional/behavioral health priorities were anxiety disorders (82 [67, 97.5]), social deficits (80 [65, 96]), and learning disabilities (79 [64.5, 96]). The top other health priorities were quality of life (82 [64, 99]), transition from pediatric to adult care (79 [67.5, 97]), and prenatal diagnosis/counseling (75.5 [60.5, 95]).

**Figure 2.**
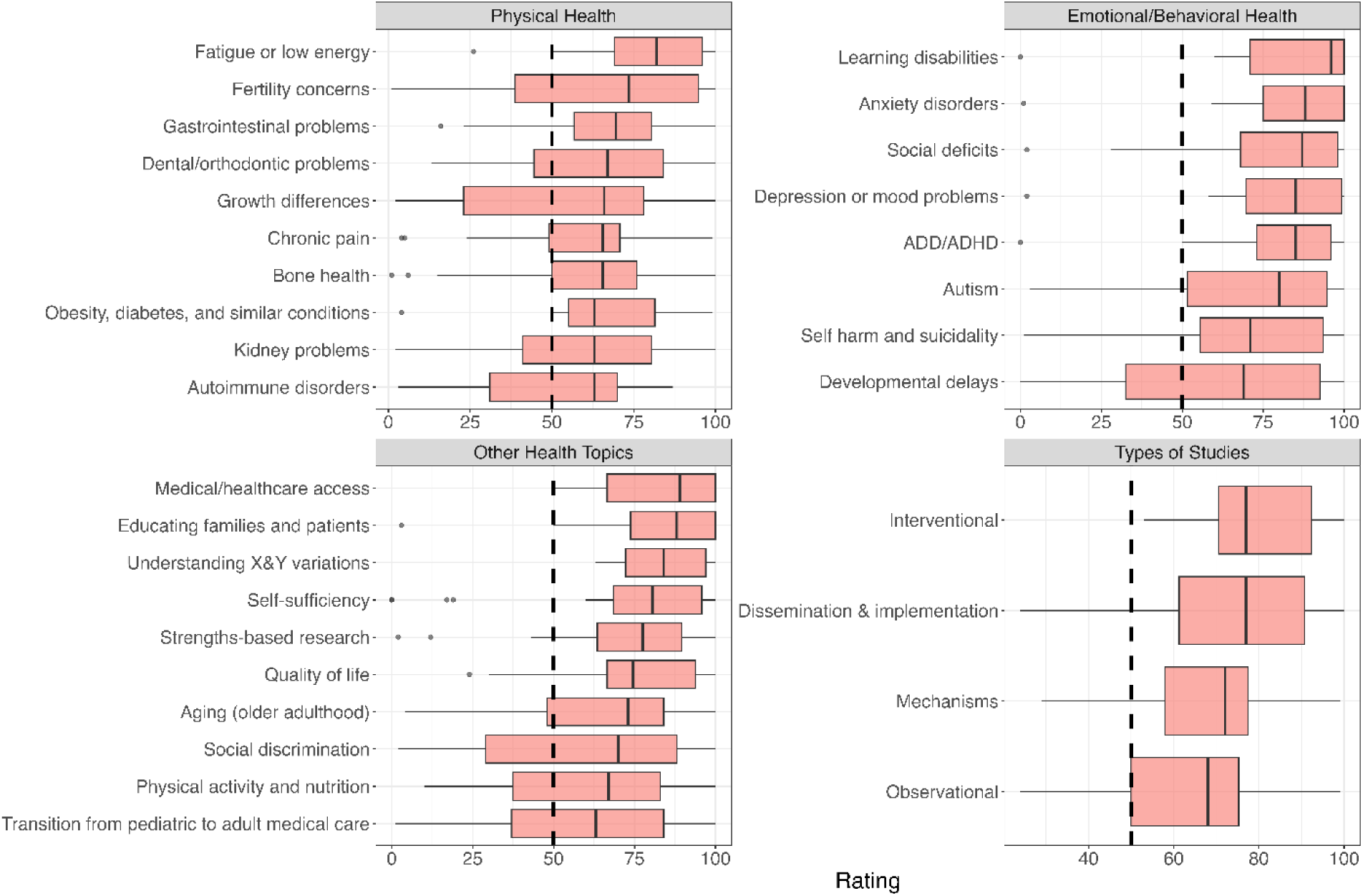
Trisomy X research priorities. Research priorities among the Triple X sample (n=17), including physical health, emotional/behavioral health, types of studies, and other health domains. Topics with a median greater than 50 are presented, capped at the top 10 topics per domain. The top physical health priorities reported were fatigue and low energy (82 [69, 96]), fertility concerns (73.5 [38.75, 94.75]), and gastrointestinal problems (69.5 [56.75, 80.5]). The top emotional/behavioral health priorities were learning disabilities (96 [71, 100]), anxiety disorders (88 [75, 100]), and social deficits (87 [68, 98]). The top other health priorities were medical or healthcare access (89 [66.5, 100]), educating families and patients (88 [73.75, 100]), and understanding X and Y variations (84 [72.25, 97]).

**Figure 3.**
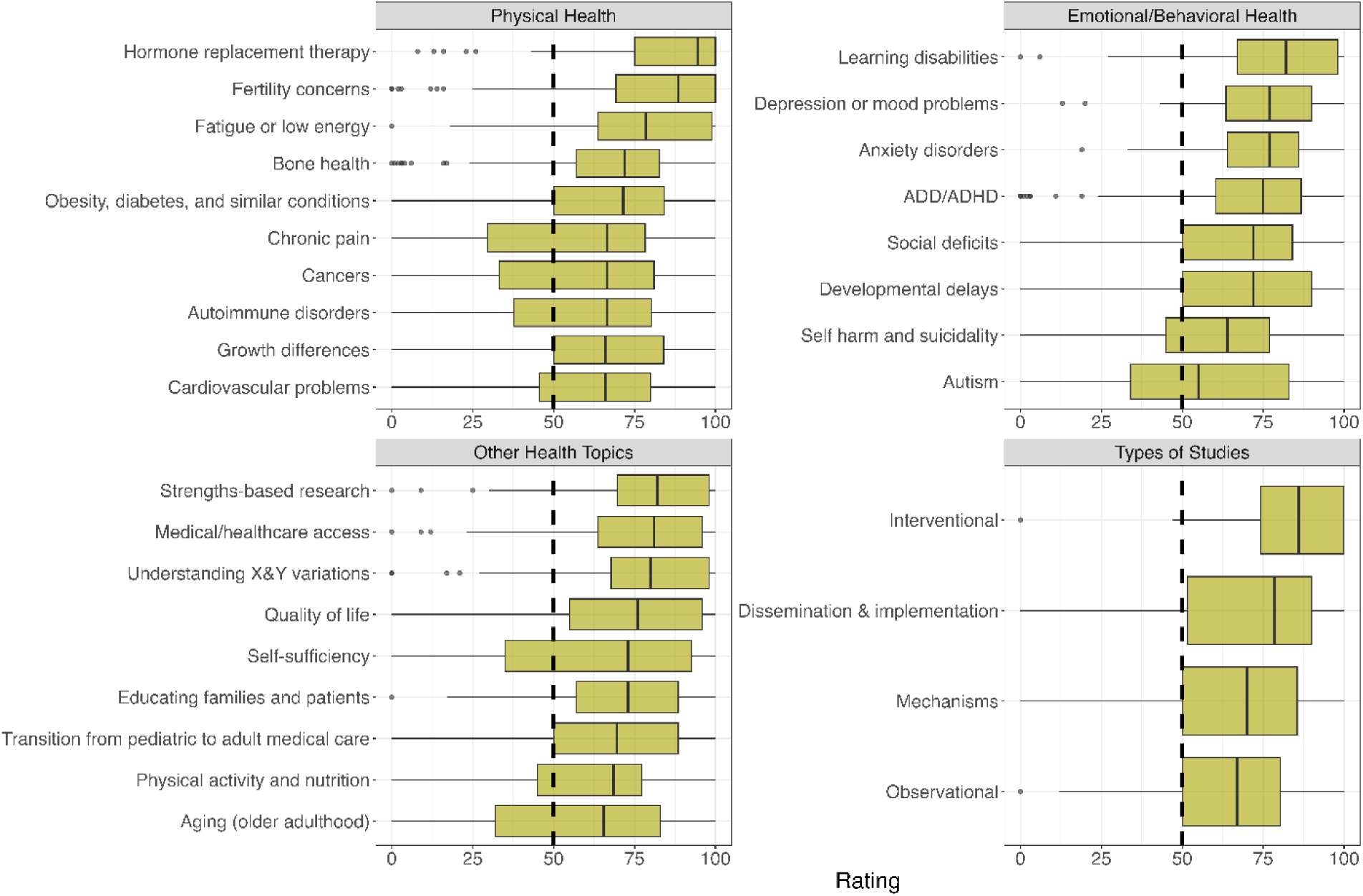
Klinefelter syndrome research priorities. Research priorities among the Klinefelter syndrome sample (n=86), including physical health, emotional/behavioral health, types of studies, and other health domains. Topics with a median greater than 50 are presented, capped at the top 10 topics per domain. The top physical health priorities reported were hormone replacement therapy (94.5 [75, 100]), fertility concerns (88.5 [69.25, 100]), and fatigue and low energy (78.5 [63.75, 99]). The top emotional/behavioral health priorities were learning disabilities (82 [67, 98]), depression or mood problems (77 [63.5, 90]), and anxiety disorders (77 [64, 86]). The top other health priorities were strengths-based research (82 [69.75, 98]), medical or healthcare access (81 [63.75, 96]), and understanding X and Y variations (80 [67.75, 98]).

**Figure 4.**
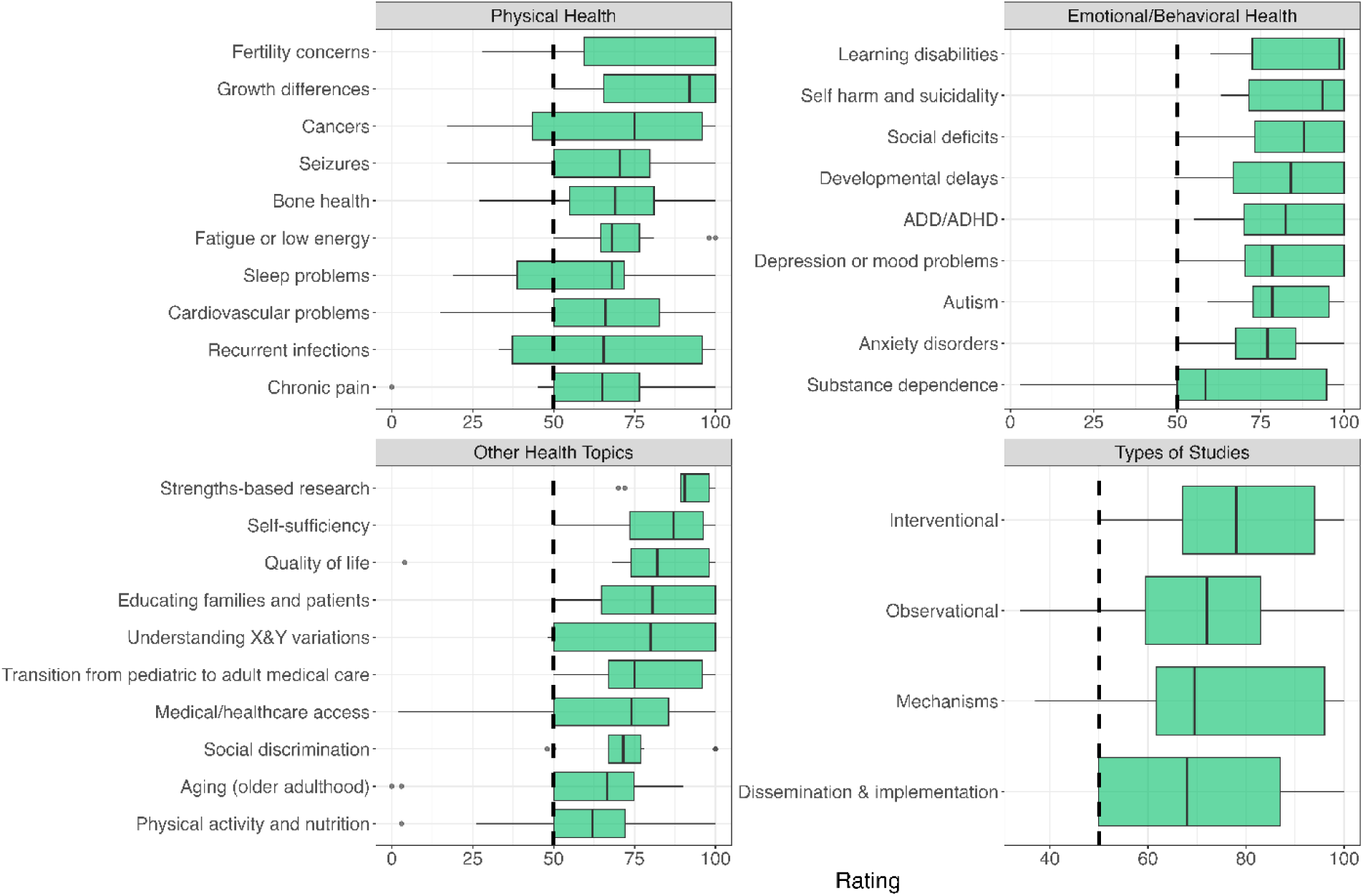
XYY research priorities. Research priorities among the XYY sample (n=12), including physical health, emotional/behavioral health, types of studies, and other health domains. Topics with a median greater than 50 are presented, capped at the top 10 topics per domain. The physical health priorities were fertility concerns (100 [59.5, 100]), growth differences (92 [65.5, 100]), and cancers (75 [43.5, 96]). The top emotional/behavioral health priorities were learning disabilities (98.5 [72.5, 100]), self-harm and suicidality (93.5 [71.5, 100]), and social deficits (88 [73.25, 100]). The top other health priorities were strengths-based research (90.5 [89.25, 98]), self-sufficiency (87 [73.5, 96.25]), and quality of life (82 [74, 98]).

**Figure 5.**
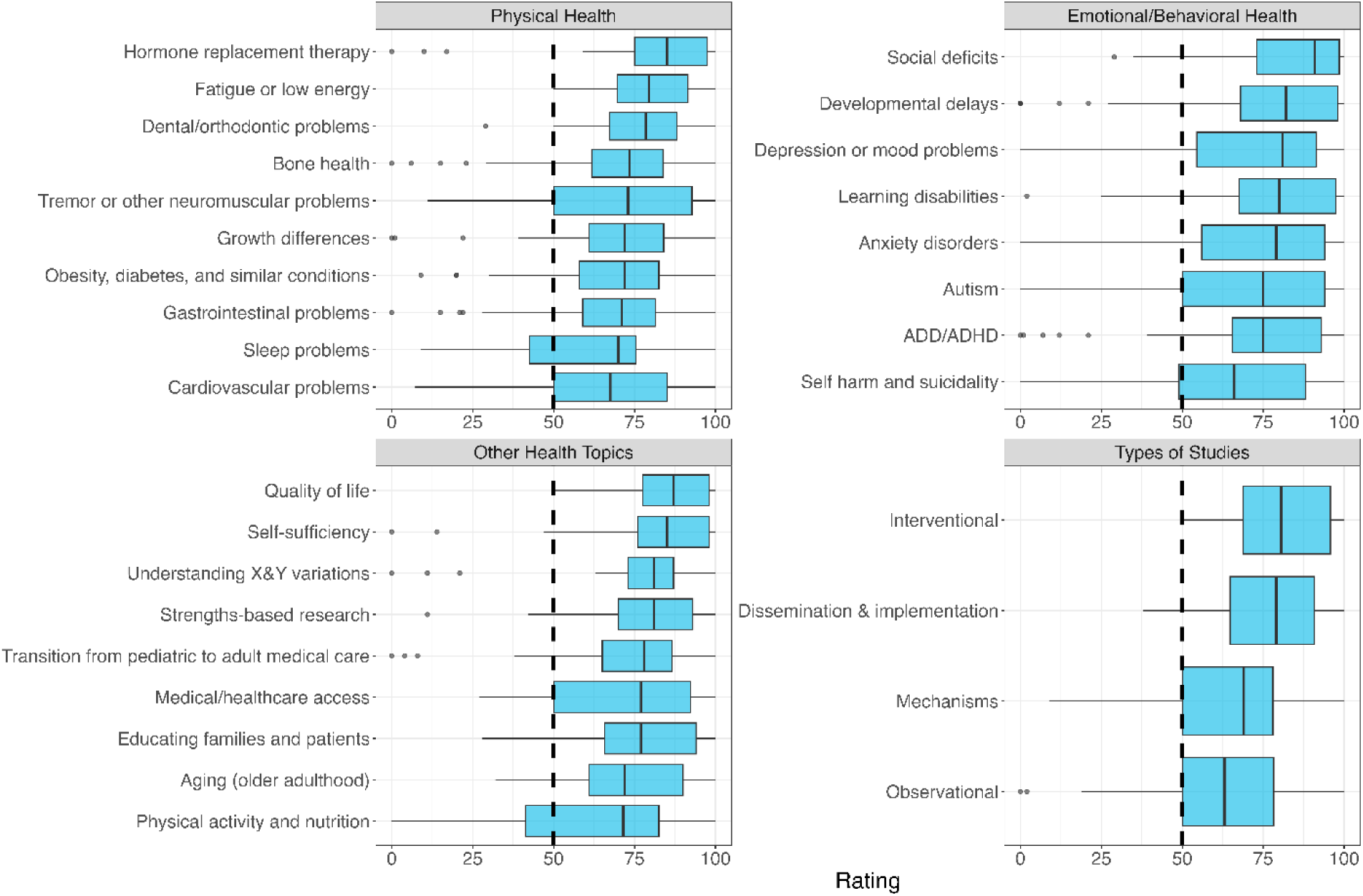
XXYY syndrome research priorities. Research priorities among the XXYY sample (n=35), including physical health, emotional/behavioral health, types of studies, and other health domains. Topics with a median greater than 50 are presented, capped at the top 10 topics per domain. The top physical health priorities reported were hormone replacement therapy (85 [75, 97.5]), fatigue or low energy (79.5 [69.75, 91.5]), and dental/orthodontic concerns (78.5 [67.25, 88]). The top emotional/behavioral health priorities were social deficits (91 [73, 98.5]), developmental delays (82 [68, 98]), and depression or mood problems (81 [54.5, 91.5]). The top other health priorities were quality of life (87 [77.5, 98]), self-sufficiency (85 [76, 98), and understanding X and Y variations (81 [73, 87]).

**Figure 6.**
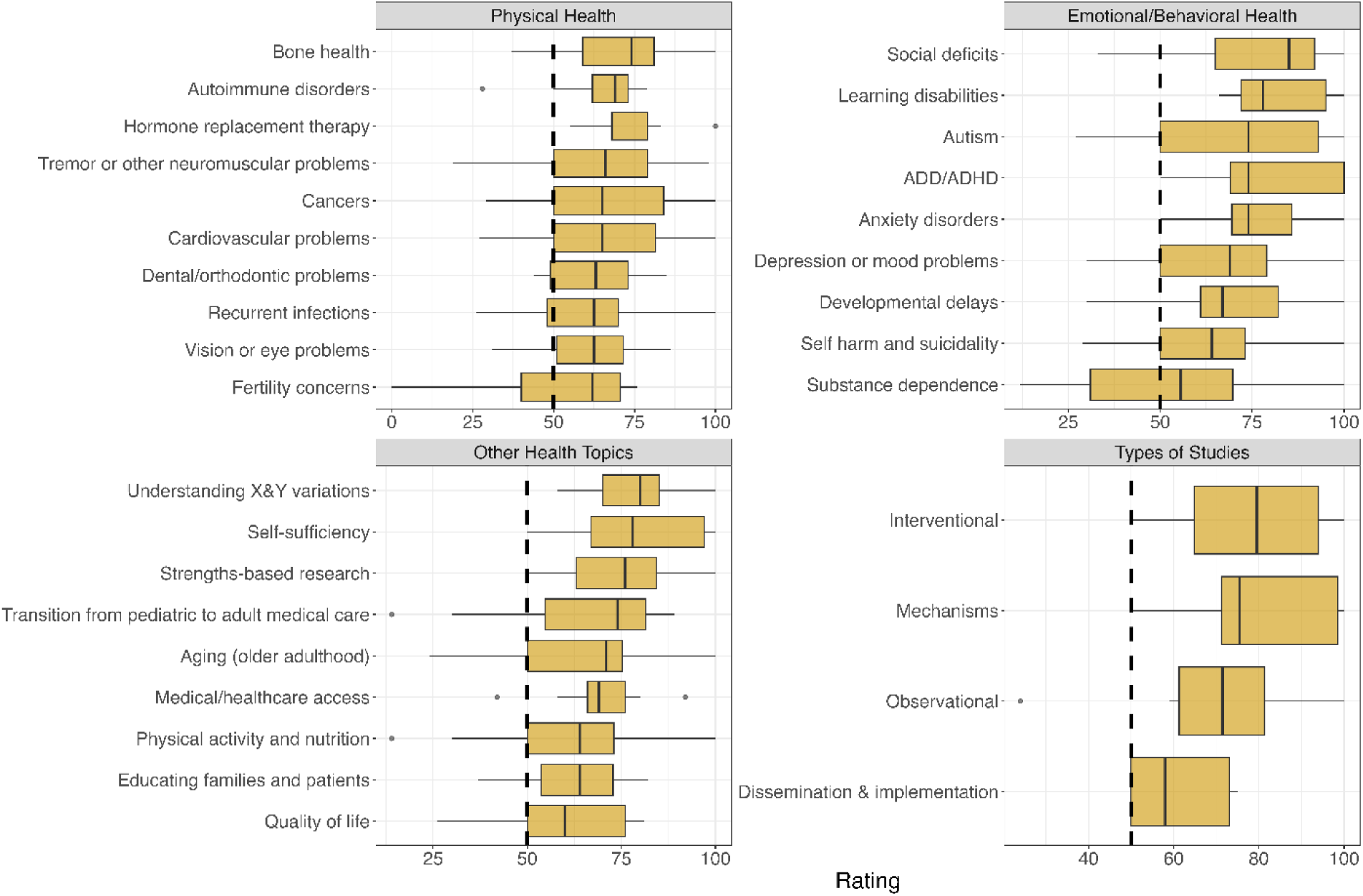
All other SCAs research priorities. Research priorities among the sample of all other SCAs (n=9), including tetrasomies and pentasomies, including physical health, emotional/behavioral health, types of studies, and other health domains. Topics with a median greater than 50 are presented, capped at the top 10 topics per domain. The top physical health priorities were bone health (74 [59, 81]), autoimmune disorders (69 [62, 73]), and hormone replacement therapy (68 [68, 79]). The top emotional/behavioral health priorities were social deficits (85 [65, 92]), learning disabilities (78 [72, 95]), and autism (74 [50, 93]). The top other health priorities were understanding X and Y variations (80 [70, 85]), self-sufficiency (78 [67, 97]), and strengths-based research (76 [63, 84.25]).

Interventional studies were identified as the highest research priority among all groups (82 [69, 97]). Additionally, all study types had a median score above 50 for all SCA groups.

### 3c. Respondent Differences

The median scores and differences for self-advocates and caregivers across all physical, emotional-behavioral, other health topics, and research study types are presented in Table 2.

Caregivers rated developmental delays as a significantly higher research priority, with a median score of 78.5 [58.75-97], substantially greater than the median of 58 [18-77] from self-advocates (p<0.001). This pattern of statistical significance was also observed for seizures where caregivers had a median score of 50 (IQR : [26.5-75]), compared to a median score of 19.5 [3-29.5] reported by self-advocates (p<0.001).

Furthermore, caregivers viewed self-sufficiency as a significantly more important research priority, with a median score of 80 [61-97], than self-advocates, who scored it at a median of 68 [31-82] (p=0.003). Similarly, caregivers rated fertility-related concerns higher, with a median of 81.5 [51.5-100], compared to a median of 33 [8-83] reported by self-advocates (p=0.004). Self-advocates rated medical/healthcare access as a higher research priority (85 [80-99]) than caregivers (76 [50-94], p=0.012).

After False Discovery Rate (FDR) adjustments, there were no significant correlations between age and any of the priority topics in the self-advocate group.

### 3d. Prioritization among SCAs

Based on hierarchical clustering analysis (Figure 7), trends in research priorities are most similar between the *TS participants* and *all other SCAs participants*, and most similar between the *Trisomy X* and *XXYY groups*. XYY respondents tended to rate research priorities higher than other groups. Across all groups, anxiety disorders, ADD/ADHD, depression or mood problems, learning disabilities, social deficits, and interventional studies are rated similarly high.

**Figure 7.**
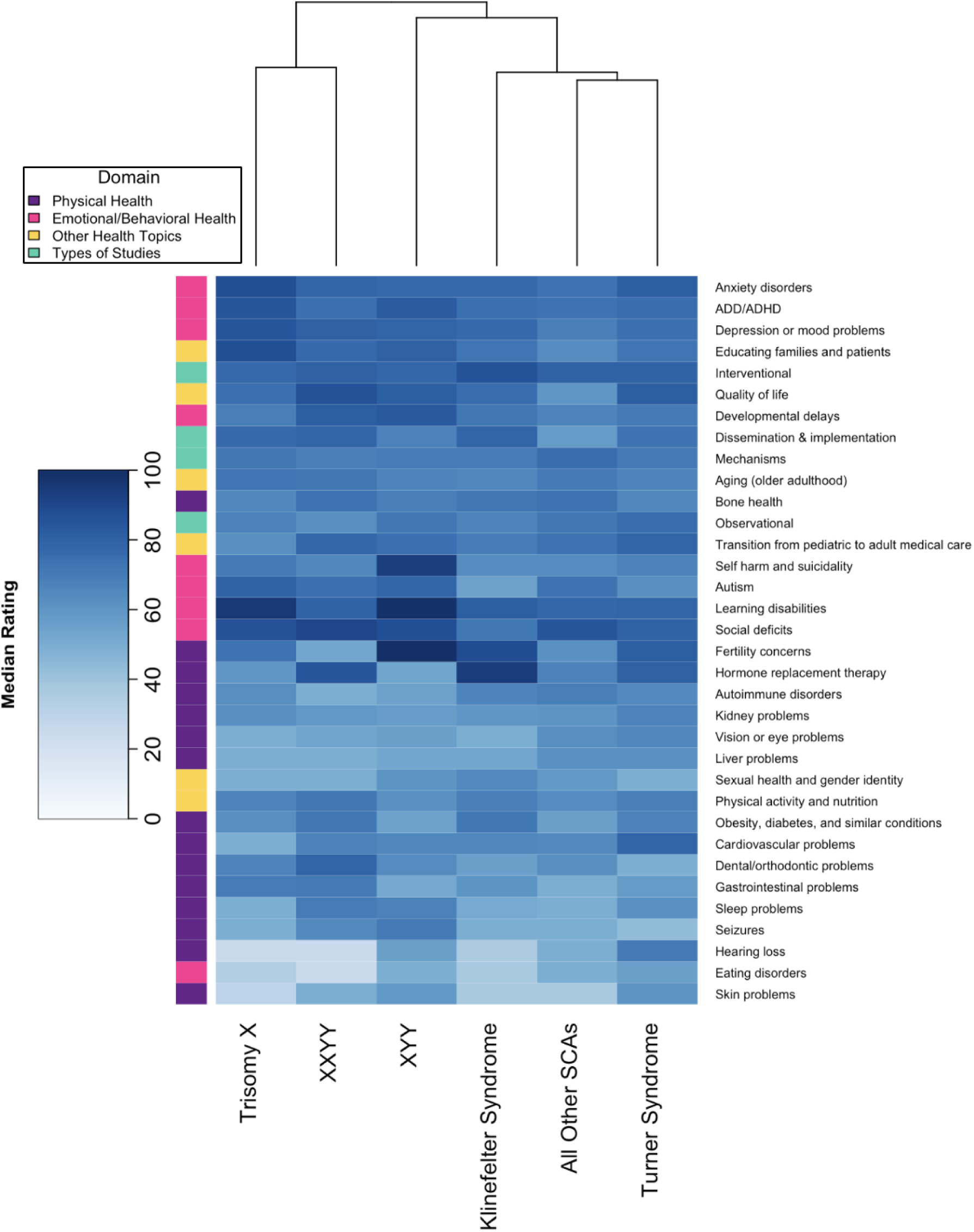
Research Priorities Heatmap of Median Ratings by SCA Diagnosis. Heatmap of median rating of research priorities by SCA, with both rows and columns sorted by hierarchical clustering to distinguish patterns between groups and priorities. Lighter colors represent lower rated priorities, with a minimum median of 25 and a maximum median of 100. Column lines represent the order of clustering, with paired lines (Trisomy X and XXYY as well as All Other SCAs and Turner Syndrome) represent SCAs that are most similar according to the clustering algorithm.

Not all groups had a sufficient sample size to compare self-advocate responses to caregiver responses. Those that could be compared are visualized in Figure 8. Based on the hierarchical analysis, caregivers of Trisomy X and XYY patients were clustered together, with their joint highest rated category being learning disabilities. Caregivers of KS and XXYY were clustered together and similarly rated hormone replacement therapy high but differed in their ratings of fertility concerns, where caregivers of KS rated this higher, possibly due to the more severe presentation of other health concerns in XXYY. Self-advocates and caregivers of TS were clustered together while self-advocates and caregivers of KS were not. Both self-advocates and caregivers of KS rated hormone replacement therapy high, however, self-advocates rated fertility concerns much lower.

**Figure 8.**
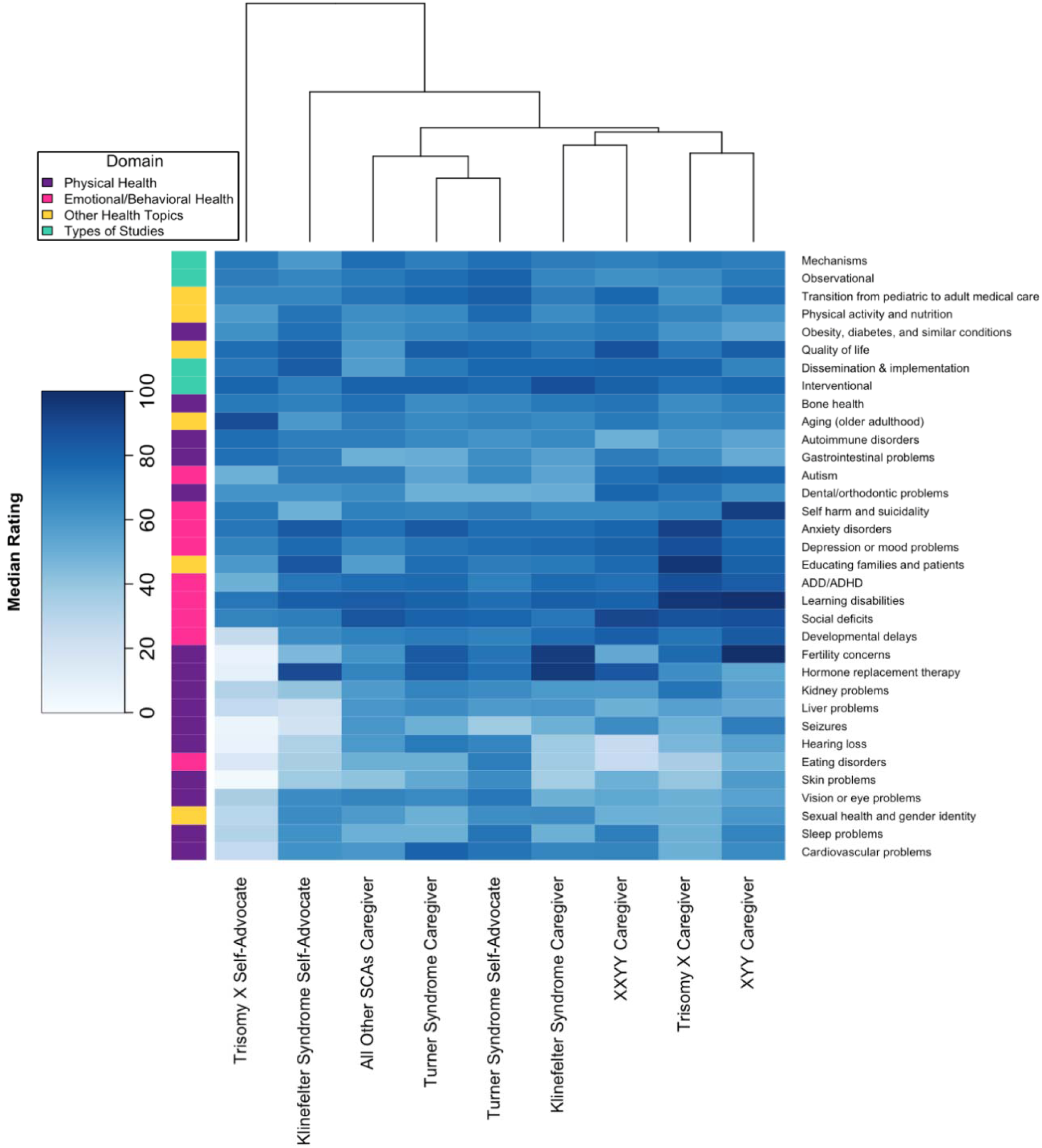
Research Priorities Heatmap of Median Ratings by SCA Diagnosis and Survey Respondent. Heatmap of median rating of research priorities by SCA and survey respondent, with both rows and columns sorted by hierarchical clustering to distinguish patterns between groups and priorities. Lighter colors represent lower rated priorities, with a minimum median of 3 and a maximum median of 100. Only groups with > 5 respondents are included in the figure. Column lines represent the order of clustering, with paired lines (such as Trisomy X and XYY Caregivers) represent groups that are most similar according to the clustering algorithm.

Survey responses were compared between participants in the INSIGHTS and GALAXY registries and are reported in supplemental table 2. After FDR adjustment for multiple comparisons, participants in the GALAXY registry rated dental/orthodontic problems significantly higher than participants in the INSIGHTS registry (65.5 [42.25, 83] and 50 [24.5, 71] respectively, p = 0.004). Participants in the INSIGHTS registry rated hearing loss (p < 0.001), cardiovascular problems (p < 0.001), vision or eye problems (p = 0.008), skin problems (p = 0.041), eating disorders (p = 0.003), transition from pediatric to adult care (p = 0.010), and observational studies (p < 0.001) significantly higher than participants in the GALAXY registry.

## 4. DISCUSSION

The current study describes results from the most detailed study of research priorities for individuals with SCAs using a community-engaged approach and are critical to addressing the needs of the SCA community. These findings provide unique insights as to what individuals with SCAs and their caregivers want research to focus on and where they feel research resources should be allocated. Importantly, intervention trials are a top priority across all SCAs, and this finding should fuel researchers to address the dearth of SCA intervention studies by developing and evaluating new interventions and treatments for multiple SCA conditions.

Prior work in the TS community suggests patients and families place importance on both physical and emotional health research^9^. These results confirmed this by showing that both physical and emotional/behavioral topics were of high importance; however, these results expand upon which specific topics were prioritized, as not all physical or emotional/behavioral topics were ranked similarly. For example, learning disabilities and anxiety disorders were of high priority in multiple SCA groups, as were fertility and hormone replacement. In all groups except KS, the top three overall priorities contained one physical health concern and 2 emotional/behavioral or other concerns. Several topics were prioritized for multiple SCA sub-groups and responders, including learning disabilities, anxiety disorders, and fatigue - the former two being well-recognized comorbidities in SCAs^13–15^, though fatigue has had minimal investigation.

These results also highlight the uniqueness of each SCA group, such as priority to research cardiovascular concerns in TS and transition to adult health care in Trisomy X. Additionally, there were some differences in priorities between self-advocates and caregivers, likely reflecting both the age differences of the individual with the SCA diagnosis as well as the perspective of the responder. We were not able to adequately assess with this cross-sectional sample how priorities may change with age of the individual with SCA, but that is a future goal of these longitudinal registries.

While there are limited data on research priorities of the SCA community, a similar approach has been applied in other rare genetic conditions. In a natural history study of Rett syndrome, caregivers were asked the top three concerns regarding their child and found that the top concerns often aligned with the child’s specific manifestation including seizures, communication concerns, and hand use^16^. Although we did not incorporate individual’s specific phenotypes, our results align generally with the idea that priorities match clinical manifestations; for example, cardiovascular concerns were high in TS and fertility concerns were high in TS and KS. Additionally, caregivers of patients with 22q deletion syndrome reported on topics they wanted to learn more about, which included practice skills, problem-solving, and communicating about the diagnosis^17^. In a qualitative study of individuals with Down syndrome, health was conceptualized as more than physical, including mental and social components such as happiness, social acceptance, and relationships^2^. While not explicit research priorities, these observations show that health priorities in other genetic conditions are unique to the phenotypes in those syndromes but also all include neurodevelopmental, emotional, and behavioral concerns.

This survey was sent to families, and only one parent or an individual completed, and thus comparison directly between caregiver and self-advocate of the same dyad was not possible. Ascertainment bias, both with individuals who receive an SCA diagnosis, those who participate in our study, and those who complete the survey, also influences the results shown here. Additionally, our sample sizes in each SCA subgroup vary widely, with especially low sample sizes in the Trisomy X, XYY, and other SCA subgroups. Therefore, comparison among the different groups is limited and results from the smaller groups should not be overly generalized to the total patient population. The sample is primarily white non-Hispanic, thus generalizing to other racial and ethnic groups as well as groups not being evaluated in SCA clinics is also limited. Future directions for this work include increasing sample size for each group, specifically focusing on Trisomy X, XYY, and the rarer SCA karyotypes (e.g., 48,XXXY and 49,XXXXY), and increasing ethnic and racial diversity of the sample.

### 4.2 Conclusions

The findings of this analysis should be used to help guide future research in subjects with SCAs. The numerous overlapping priorities among SCA subgroups may enable researchers to conduct transdiagnostic group studies with increased sample size and statistical power. At the same time, we also identified unique features that distinguish the SCA subgroups, and careful considerations should be made to avoid over-generalizing among the diagnoses. Results highlight the importance of a multidisciplinary focus for addressing research needs of the SCA population. To be responsive to SCA community priorities, SCA researchers are encouraged to collaborate and target research efforts toward clinical trial readiness and interventional studies for both medical and psychological manifestations of SCA conditions.

## Data Availability

All data produced in the present study are available upon reasonable request to the authors

